# Provider attitudes toward precision medicine for brain arteriovenous malformations: a survey study

**DOI:** 10.64898/2026.11.26.26369091

**Authors:** Andrew T. Hale, Jonah Keller, Caroline Davies, Neel H. Mehta, Christopher Sollenberger, Trisha McElhaugh, Shay Overstone, Patricia L. Musolino, Denise H. Adams, Brandon G. Rocque, Phillip B. Storm, Edward R. Smith, Kristopher T. Kahle, Adam J. Kundishora

## Abstract

**Objective:** Somatic *KRAS* and *BRAF* mutations drive sporadic brain arteriovenous malformations (bAVMs) through endothelial RAS-MAPK hyperactivation, suggesting opportunities for molecularly targeted treatment. We assessed cerebrovascular specialists’ attitudes toward precision therapies to guide patient selection and inform clinical trial design.

**Methods:** A cross-sectional survey comprising 35 general questions and 12 case vignettes was distributed to 70 cerebrovascular specialists in October-December 2025. Mixed-effects logistic regression with a random intercept for each respondent assessed endorsement of any molecular therapy by Spetzler-Martin (SM) grade. Secondary analyses assessed therapy type, treatment timing, rupture status, and age.

**Results:** Forty-eight specialists responded (69%; 85% male; 54% full professors); 90% considered drug therapy feasible and 92% supported randomized trials of drug therapy. Among 436 vignette observations from 39 respondents, molecular therapy endorsement increased from 69% at SM grade I to 92% at grade V (odds ratio [OR] per grade, 10.41; 95% CI, 4.10-26.42; *P* < .001). Grade associations were significant for drug therapy (OR, 6.74; 95% CI, 3.21-14.14) and gene editing (OR, 2.51; 95% CI, 1.61-3.92; both *P* < .001). First-line drug therapy endorsement increased from 15% to 75% across these grades. Prior rupture (OR, 2.44; 95% CI, 1.70-3.50) and age <20 years (OR, 3.47; 95% CI, 2.26-5.32) independently increased endorsement (both *P* < .001). Findings remained robust across four prespecified sensitivity analyses.

**Conclusions:** Cerebrovascular specialists broadly supported molecular therapies for bAVMs, particularly for higher-grade lesions, ruptured presentations, and younger patients. These findings support precision medicine trials prioritizing high-grade bAVMs and development of endovascular-enabled molecular diagnostics and biomarkers.

## Introduction

Treatment of brain arteriovenous malformations (bAVM) currently involves limitation to neurosurgical resection, stereotactic radiosurgery (SRS), or adjunctive endovascular embolization, currently dictated by several factors, with each being associated with specific procedural morbidity and mortality risk. However, recent genetic discoveries may have application in the care of various bAVMs as they have identified recurrent, actionable somatic mutations in *KRAS*, and less commonly *BRAF*, in 50-80% of sporadic bAVMs^1–5^. These mutations localize to endothelial cells and hyperactivate RAS/MAPK signaling.

Although these mutations are common across cancer types and are active targets for drug development, including FDA-approved pharmacotherapies^6^, no medical therapies for bAVMs currently exist in this treatment category. However, more recently, studies have begun to address this gap through pharmacologic and gene editing strategies in preclinical models. Together, their findings suggest that molecularly targeted therapies may be viable, but it remains unclear which bAVMs should be treated in these ways and what provider attitudes may be toward acceptance and utilization of these potential treatments that illustrate a paradigm shift.

To explore this, we conducted a cross-sectional survey of multidisciplinary cerebrovascular providers to assess attitudes toward implementing such precision medicine for bAVMs^13^. We established provider experience with endovascular-enabled molecular diagnostics and evaluations of pharmacologic and gene editing therapies. The survey therefore included a systematic, case vignette-based participant appraisal of genotype-directed treatments involving timing (first-line, neoadjuvant, adjuvant), according to rupture history and patient age based on the Spetzler-Martin (SM) and Lawton-Young (LY) grading scales^14,15^. These findings could provide empirical support for the design and implementation of genotype-stratified clinical trials in sporadic bAVMs.

## Methods

### Study design and reporting standards

A cross-sectional survey to quantify provider perspectives on precision medicine treatments (drug and gene editing) for treatment of bAVMs was developed, involving general questions as well as case-based vignettes addressing components of the SM and LY scores^14,15^ that included medical illustrations by Laura Roy.

The study was conducted and reported in accordance with STROBE guidelines for cross-sectional studies, AAPOR best practices for survey research, and the CHERRIES checklist for web-based surveys. Cerebrovascular providers, including neurosurgeons and endovascular specialists across the age spectrum, were assessed. Participant demographics (age, gender, professional background, and experience with molecular therapies and endovascular diagnostics) were collected.

#### Study population, recruitment, and data collection

Eligible specialty participants were identified through professional networks and specialty-based distribution lists, including current and past leaders of regional, national, and international academic societies with professional focus on bAVM care. Invitations were distributed by email between October-December 2025. Three reminders were sent at two-week intervals; non-respondents were contacted by personalized text or phone call. The survey was administered electronically without financial or material incentives.

### Survey instrument

The survey included 35 general questions on perspectives toward precision medicine for bAVMs, with a focus on drug and gene-editing treatment, and 12 case scenarios (**Supplementary Note 1**). The instrument was developed by the study team and was not derived from any previously validated questionnaire. Prior to dissemination, it was reviewed and responded to by all authors; authors’ responses were excluded from analysis.

In addition, 12 bAVM case vignettes were constructed and included to provide a standardized framework for evaluating patient candidacy, interpreting outcomes, and guiding clinical trial design. Each vignette included six structured questions on treatment preferences based on bAVM size, anatomical location, rupture history, and patient age, systematically addressing the SM and, in part, the LY scores^14,15^. Although the LY score has been externally validated, the subjective determination of “nidus compactness” has not undergone formal interrater reliability testing and was therefore omitted. Vignettes varied SM grade and anatomical location. Participants indicated treatment recommendations (drug and/or gene editing) and intervention timing (first-line, adjunctive, salvage). In addition, “No Therapy” was available as a response. The survey took approximately twenty minutes to complete.

### Data management and confidentiality

Survey data were protected by being stored in REDCap, which also ensured confidentiality.

### Statistical analysis

Responses were aggregated and analyzed using descriptive statistics. A potential sample of 40 respondents each evaluating 12 vignettes yields 480 observations, sufficient to estimate an odds ratio (OR) > 3.0 with 80% power at α = 0.05 in a mixed-effects model. For each of 10 binary therapy endorsement outcomes (five primary, five secondary), mixed-effects logistic regression models of the form: outcome ∼ SM grade (continuous, one through five) + (1 | respondent), using the glmer function (family = binomial, bobyqa optimizer) in lme4 were utilized. The SM grade exponentiated coefficient yields the OR per one-unit increase in SM grade, with a random intercept accounting for repeated measures. Observed endorsement proportions with exact binomial 95% confidence intervals (CI) were computed at each SM grade.

To quantify variance attributable to inter-provider differences, Intraclass correlation coefficients (ICC) were calculated using the performance package. Model convergence and singularity were assessed programmatically. P-values were adjusted using the Benjamini-Hochberg false discovery rate (FDR) method: Bonferroni-corrected values are also reported.

Four prespecified sensitivity assesses robustness: (S1) adjustment for provider training subspecialty, (S2) adjustment for AVM anatomical location, (S3) restriction to complete-case respondents, and (S4) dichotomization of SM grade (I-II vs III-IV).

### Missing data and non-response bias

Respondent characteristics were compared across all demographic variables in Table 1 using Chi-Squared tests. Missing data were minimal; 36 of 39 case-based respondents completed all 12 scenarios. The 3 partial completers contributed only their completed scenarios; incomplete scenarios were excluded rather than imputed.

**Table 1.** Participant demographics and characteristics of respondents (n = 48) and non-respondents (n = 22) are shown across all available characteristics. *P* values reflect chi-squared or Fisher’s exact tests comparing respondents versus non-respondents.

| **Characteristic** | **Responders<br>(n = 48)** | **Non-responders<br>(n = 22)** | ***P* Value** |
|---|---|---|---|
| **Overall response rate** | 48/70 (69%) | 22/70 (31%) |  |
| **Age** |  |  | 0.33 |
| 35–44 | 17 | 7 |  |
| 45–54 | 12 | 6 |  |
| 55–64 | 15 | 4 |  |
| 65–74 | 4 | 5 |  |
| **Sex** |  |  | 0.66 |
| Male | 41 | 18 |  |
| Female | 6 | 4 |  |
| Prefer not to answer | 1 | 0 |  |
| **Location** |  |  | 0.94 |
| US-based | 44 | 21 |  |
| Non-US based | 4 | 1 |  |
| **Practice training background** |  |  | 0.66 |
| Adult – Open & endovascular | 20 | 12 |  |
| Adult – Open vascular only | 12 | 4 |  |
| Adult – Endovascular only | 4 | 3 |  |
| Pediatric – Open vascular | 10 | 2 |  |
| Pediatric – Open & endovascular | 1 | 0 |  |
| Pediatric – Endovascular only | 1 | 1 |  |
| **Practice type** |  |  | 0.22 |
| Adult only | 9 | 6 |  |
| Pediatrics only | 9 | 1 |  |
| Combined (majority adult) | 27 | 15 |  |
| Combined (majority pediatric) | 3 | 0 |  |
| **Practice setting** |  |  | 0.25 |
| Adults-only hospital | 8 | 6 |  |
| Stand-alone children's hospital | 10 | 2 |  |
| Combined hospital | 26 | 14 |  |
| Other | 4 | 0 |  |
| **Board certified** |  |  | 0.33 |
| No, not board eligible | 1 | 0 |  |
| No, board eligible | 5 | 5 |  |
| Yes | 42 | 17 |  |
| **Academic rank** |  |  | 0.38 |
| Assistant Professor | 9 | 3 |  |
| Associate Professor | 11 | 9 |  |
| Full Professor | 26 | 10 |  |
| Not applicable | 2 | 0 |  |
| **Highest research training** |  |  |  |
| No formal training | 15 | - |  |
| Funded fellowship (non-degree) | 13 | - |  |
| Master's degree | 4 | - |  |
| Doctorate (PhD) | 13 | - |  |
| Other | 3 | - |  |
| **Research grants awarded\*** |  |  |  |
| No | 3 | - |  |
| NIH or equivalent – Principal Investigator | 17 | - |  |
| NIH or equivalent – Co-investigator | 20 | - |  |
| Foundation grant – Principal Investigator | 27 | - |  |
| Foundation grant – Co-investigator | 15 | - |  |
| Institutional award – Principal Investigator | 24 | - |  |
| Institutional award – Co-investigator | 15 | - |  |
| **Heard 'endoluminal biopsy' before?** |  |  |  |
| No | 4 | - |  |
| Yes | 44 | - |  |
| **Heard 'liquid biopsy' before?** |  |  |  |
| No | 2 | - |  |
| Yes | 46 | - |  |

*P values from chi-squared or Fisher’s exact test comparing respondent’s vs non-respondents. Indented rows display subcategory count. Characteristics describing research training, grants, and biopsy familiarity were not compared between groups due to incomplete data availability. Respondents could select multiple research grant subcategories.*

### Ethics

The study was approved by [BLINDED FOR REVIEW] Institutional Review Board (IRB). All participants provided informed consent. Confidentiality was maintained by aggregating responses and removing identifying information. Participants could withdraw at any time.

### Software and code availability

Statistical analyses were performed using R version 4.5.1 (R Foundation for Statistical Computing). Mixed-effects models were fit with lme4 (v1.1.37); model diagnostics used the performance package (v0.15.2); figures were generated with ggplot2 (v4.0.0); tables with kableExtra. All code is available at [BLINDED FOR REVIEW].

## Results

### Survey Responses and Characteristics

Of 70 invited cerebrovascular specialists, 48 responded (69% response rate; **Table 1**). The cohort was predominantly male (41/48 [85%]), with 26/48 (54%) holding the rank of full professor and 30/48 (62%) maintaining a mixed adult-pediatric practice. Among respondents, 25/48 (52%) had endovascular training; 5 with exclusively endovascular training were excluded from case-scenario analyses and 4 additional respondents did not complete any case scenarios, leaving 39 respondents contributing 436 observations. Comparison of responders and non-responders across demographic variables revealed no significant differences (all *P* > 0.22; **Table 1**).

### General Attitudes Toward Precision Medicine

Overall, 43/48 respondents (90%) believed in the feasibility of drug therapy for bAVMs, while 28/48 (58%) considered gene editing feasible (**eTable 1**). Asked whether they would consider offering these therapies for any indication, 40/48 (83%) endorsed drug therapy and 25/48 (52%) endorsed gene editing, with enthusiasm highest in salvage settings: 43/48 (90%) for drug therapy and 30/48 (62%) for gene editing. A strong majority supported randomized clinical trials: 44/48 (92%) for drug therapy and 32/48 (67%) for gene editing. Additionally, 40/48 (83%) indicated that multidisciplinary committee adjudication should be required for drug therapy and 38/48 (79%) for gene editing.

### Endovascular Diagnostic Attitudes

Among the 25 respondents with endovascular training (1 did not answer the training question and was excluded), 12/25 (48%) had previously performed endoluminal biopsy and 13/25 (52%) had performed liquid biopsy for any indication (**eTable 2**). For bAVMs specifically, only 7/25 (28%) had performed endoluminal biopsy and 5/25 (20%) had performed liquid biopsy. Looking forward, 24/25 (96%) would consider incorporating endoluminal biopsy and 25/25 (100%) would consider liquid biopsy into their practice, while 18/25 (72%) expressed interest in obtaining molecular data as part of standard endovascular care.

### Association of Spetzler-Martin Grade With Precision Therapy Endorsement

Case vignettes were designed according to the SM grading scale (**Figure 1**) to provide a standardized framework for evaluating patient candidacy and clinical trial design. Across the 12 SM-graded case vignettes (**Figure 2**), the likelihood of recommending any molecular therapy increased monotonically, rising from 69% at SM I to 92% at SM V (*P* < .001; **Figure 3**; **eTables 3-4**). The odds of endorsing any molecular therapy increased significantly per one-unit increase in SM grade (OR, 10.41; 95% CI, 4.10–26.42; *P* < .001). This effect was independently driven by drug therapy (OR, 6.74; 95% CI, 3.21–14.14; *P* < .001) and gene editing (OR, 2.51; 95% CI, 1.61–3.92; *P* < .001). Among secondary outcomes, first-line drug therapy (OR, 4.21; 95% CI, 2.98–5.96; *P* < .001) and first-line gene editing (OR, 3.38; 95% CI, 2.28–5.00; *P* < .001) demonstrated the largest effects, indicating that the strongest endorsement per SM grade unit was for primary use rather than as adjuvant or salvage options. Respondents’ opinion shifted away from favoring salvage use toward first-line use with increasing SM grade: for drug therapy, first-line endorsement rose from 15% at SM I to 75% at SM V while salvage endorsement declined from 55% to 28%; a similar pattern was observed for gene editing (first line: from initially 11% to ultimately 44%; salvage: from 39% to 22%). Endorsement of “no therapy” declined from 31% at SM I to 8% at SM V (OR, 0.14; 95% CI, 0.07–0.30; *P* < .001). These expressed endorsement proportions confirmed a monotonic dose–response relationship across all therapy categories (**Figure 4**; **eFigure 1-2**).

**Figure 1.**
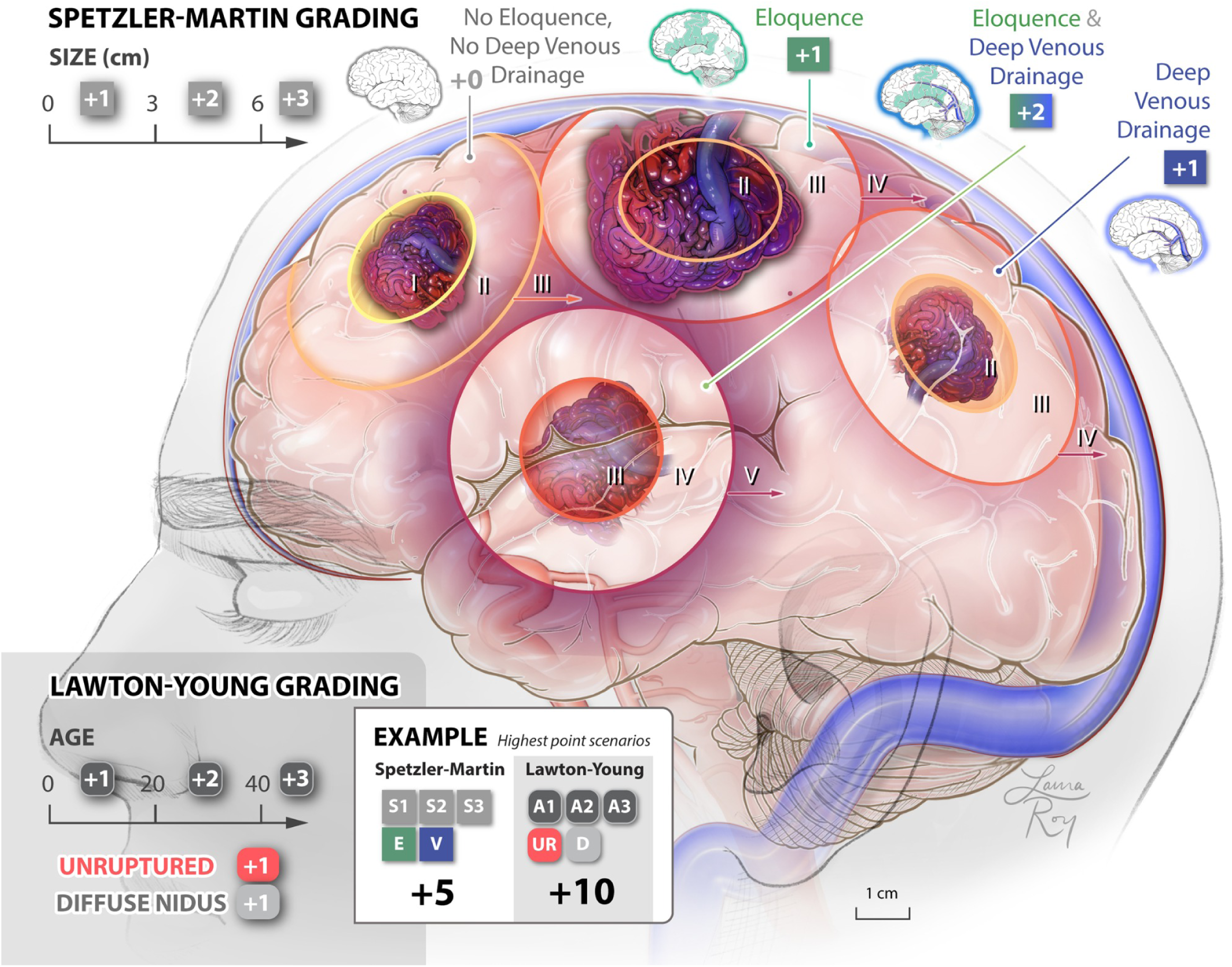
Spetzler–Martin and Lawton–Young brain AVM grading scales. The Spetzler-Martin scale grades bAVMs I-V based on nidus size (< 3 cm = 1 point; 3-6cm = 2 points; > 6 cm = 3 points), eloquence of adjacent brain (non-eloquent = 0; eloquent = 1), and venous drainage (superficial = 0; deep = 1). The Lawton-Young supplementary scale assigns additional points for patient age, unruptured presentation, and a diffuse nidus. The combined scale ranges from 1 to 10. Illustration by Laura Roy Illustrations.

**Figure 2.**
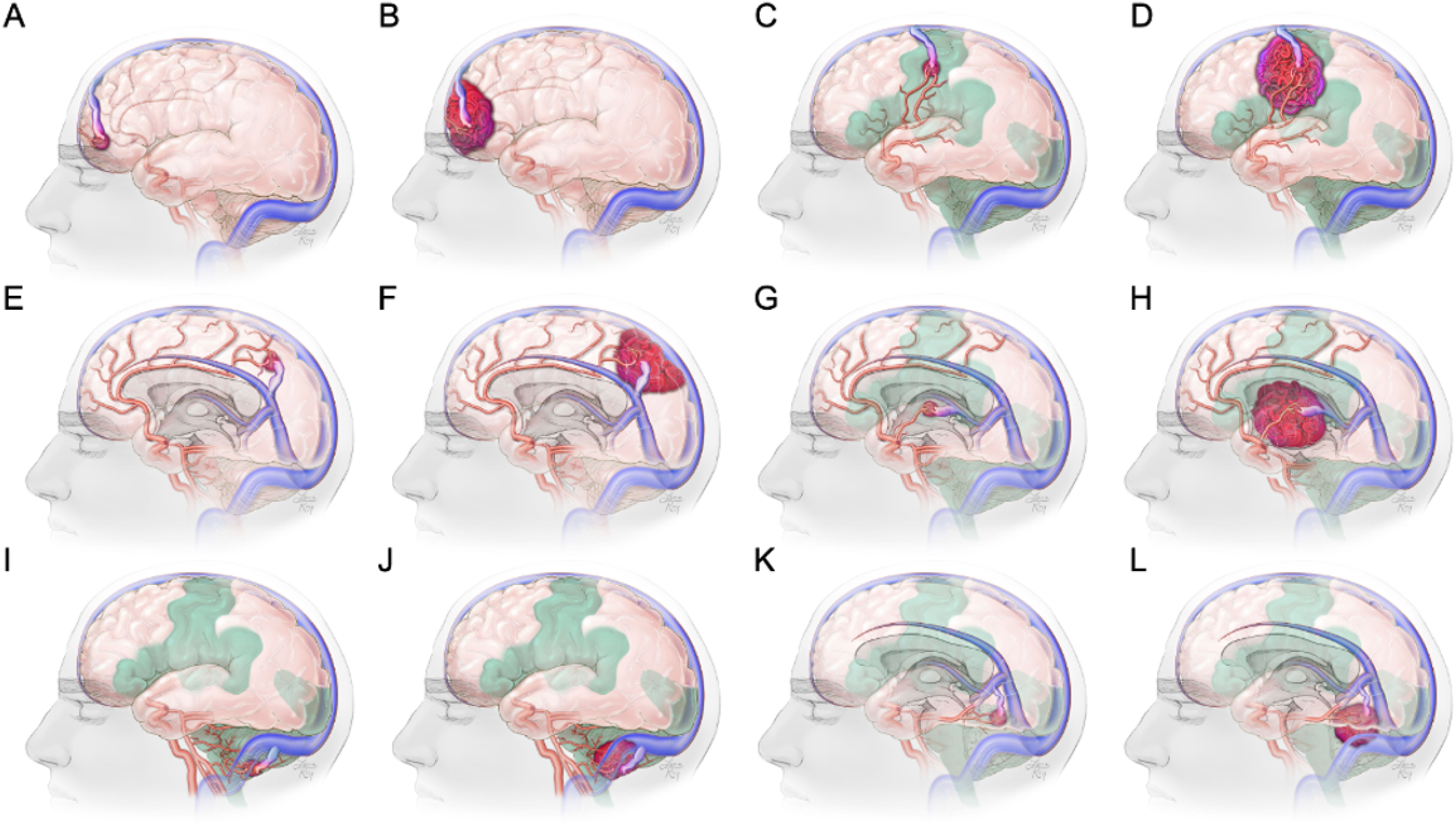
Brain AVM case vignettes for consideration of molecular targeted therapies. Twelve cases span SM grades I–V across six anatomical locations with varying nidus size, eloquence, and venous drainage patterns. A) Case 1: Spetzler-Martin grade I AVM in the frontal pole, < 3 cm with anterior cerebral artery feeders, and superficial drainage to the superior sagittal sinus. (B) Case 2: Spetzler-Martin grade III AVM in the frontal pole, > 6 cm in size, with anterior cerebral artery feeders, and superficial drainage to the superior sagittal sinus. (C) Case 3: Spetzler-Martin grade II AVM in the eloquent motor/somatosensory cortex, < 3 cm in size, with middle cerebral artery feeders, and superficial drainage to the superior sagittal sinus. (D) Case 4: Spetzler-Martin grade IV AVM in the eloquent motor/somatosensory cortex, > 6 cm in size, with middle cerebral artery feeders, and superficial drainage to the superior sagittal sinus. (E) Case 5: Spetzler-Martin grade II AVM in the non-eloquent precuneus, < 3 cm in size, with anterior cerebral and middle cerebral feeders, and deep venous drainage. (F) Case 6: Spetzler-Martin grade IV AVM in the non-eloquent precuneus, > 6 cm in size, with anterior cerebral and middle cerebral feeders, and deep venous drainage. (G) Case 7: Spetzler-Martin grade III AVM in the eloquent thalamus/basal ganglia, < 3 cm in size, with middle cerebral artery perforator feeders, and deep venous drainage. (H) Case 8: Spetzler-Martin grade V AVM in the eloquent thalamus/basal ganglia, > 6 cm in size, with middle cerebral artery perforator feeders, and deep venous drainage. (I) Case 9: Spetzler-Martin grade I AVM in the non-eloquent cerebellar hemisphere, < 3 cm in size, with posterior inferior cerebellar artery perforator feeders, and superficial venous drainage. (J) Case 10: Spetzler-Martin grade II AVM in the non-eloquent cerebellar hemisphere, 3-6 cm in size, with posterior inferior cerebellar artery perforator feeders, and superficial venous drainage. (K) Case 11: Spetzler-Martin grade III AVM in the eloquent vermis, < 3 cm in size, with posterior inferior cerebellar artery perforator feeders, and deep venous drainage. (L) Case 12: Spetzler-Martin grade IV AVM in the eloquent vermis, 3-6 cm in size, with posterior inferior cerebellar artery perforator feeders, and deep venous drainage.

**Figure 3.**
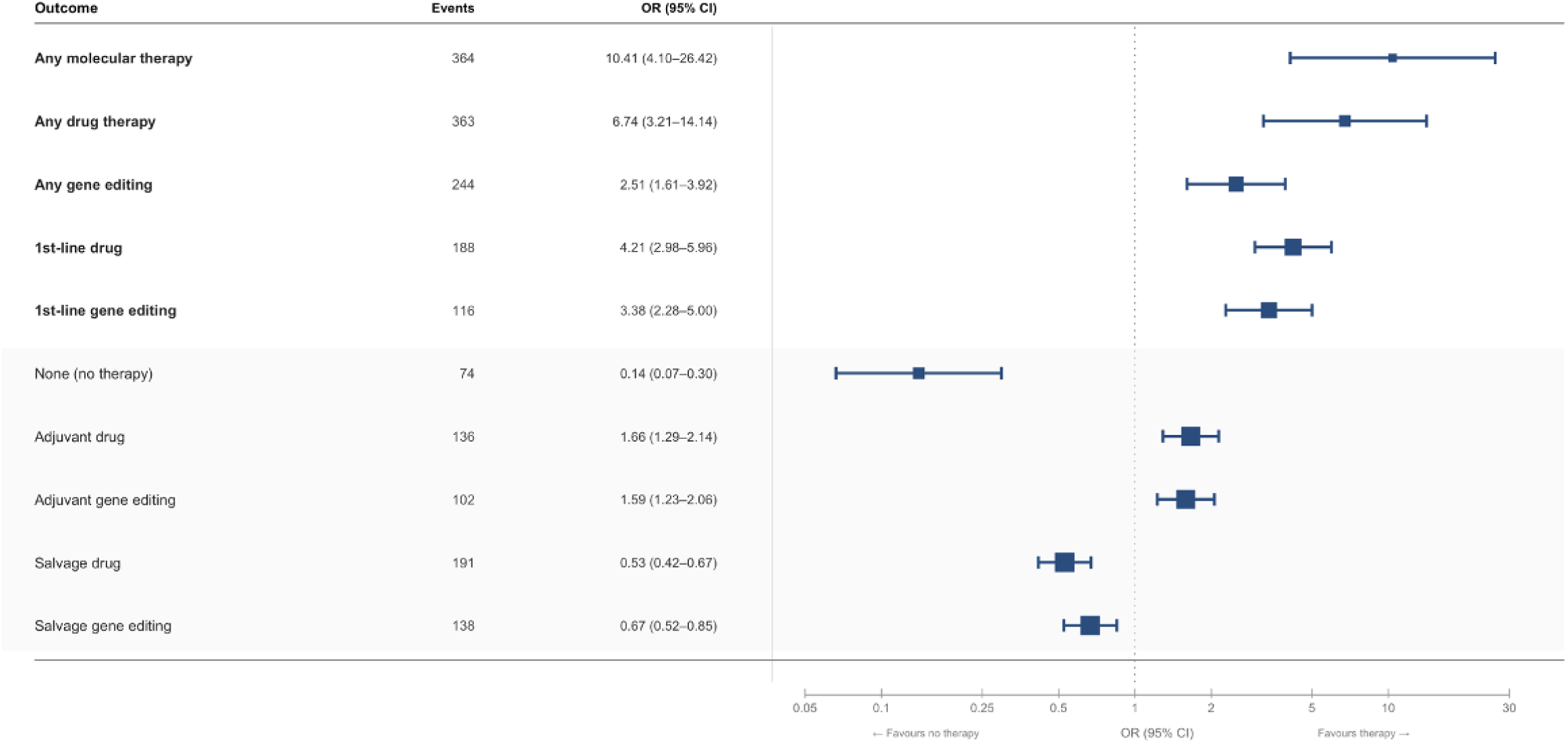
Forest plot of mixed-effects logistic regression results. Each point represents the odds ratio (OR) per Spetzler-Martin grade unit increase in treatment recommendation probability, estimated from a model with a random intercept for respondent to account for repeated measures. Each respondent evaluated up to 12 case vignettes. Horizontal lines indicate 95% confidence intervals (CI). The five primary outcomes (any molecular targeted therapy, any drug therapy, any gene editing, first-line drug therapy, and first-line gene editing) are shown in bold. Five secondary outcomes (no therapy, adjuvant drug, adjuvant gene editing, salvage drug, and salvage gene editing) are in the grey shaded region below. All ORs are per 1 unit increase in SM grade; ORs > 1 indicate increasing recommendation probability with increasing SM grade. ORs < 1 indicate decreasing probability. All 10 models remained statistically significant after Benjamini-Hochberg false discovery rate (FDR) correction (all adjusted P < .001). See eTable 4 for full model results including event counts.

**Figure 4.**
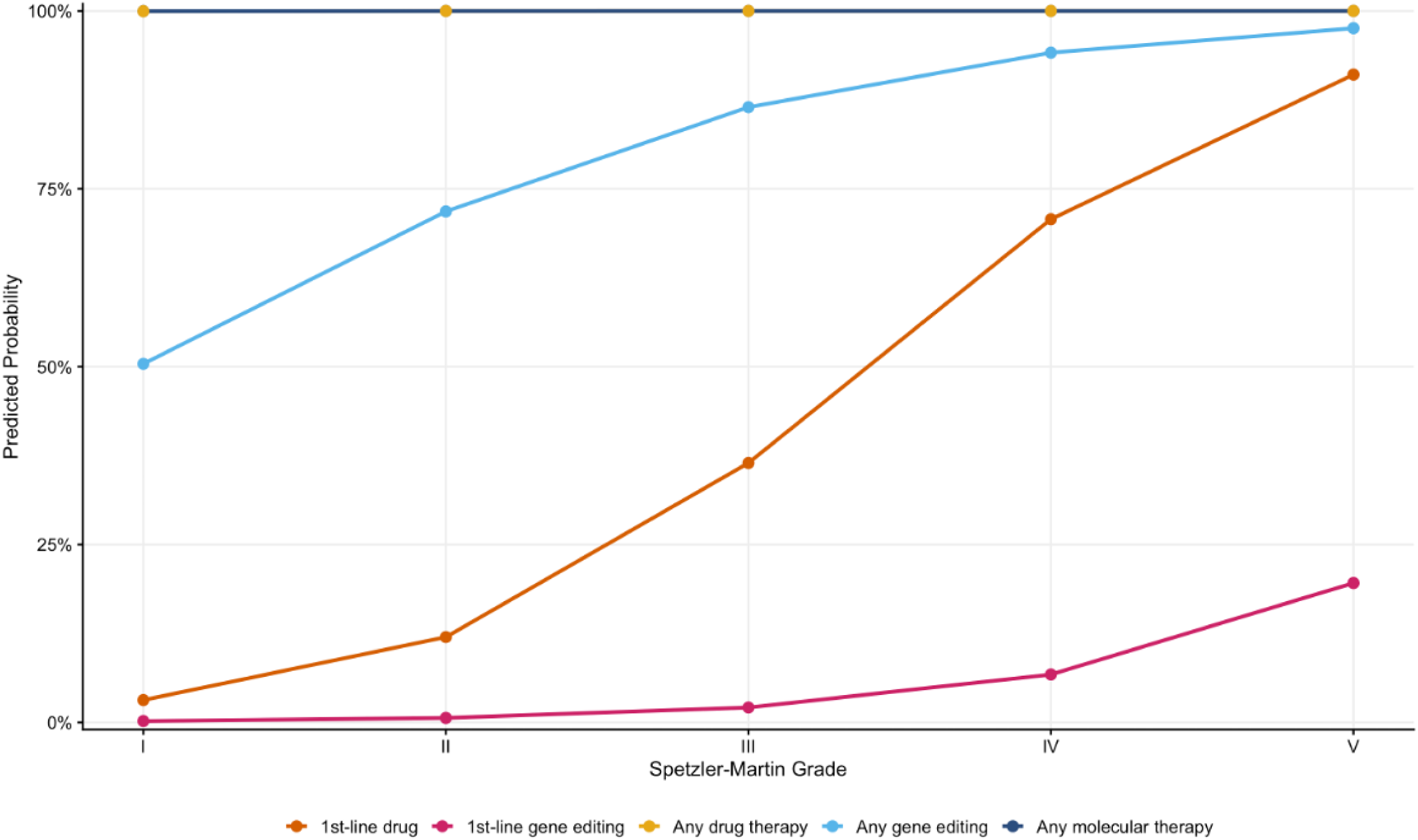
Observed proportion of therapy endorsement by Spetzler–Martin grade. Points represent the observed recommendation proportion (y-axis) across all respondents for each SM grade I-V (x-axis). Lines connect observations across grades to illustrate the dose-response relationship. Colors distinguish therapy outcomes by category. SM grades I-V correspond to 39, 39, 117, 117, and 39 respondent-observations, respectively, after exclusion of endovascular-only training respondents.

All 10 models met the stringent FDR correction (all adjusted *P* < .001). ICCs ranged from 0.70 to 0.98 (**eTable 4**), indicating that inter-provider variability – rather than case-level angioarchitectural features – accounted for the most variance in therapy recommendations. The high upper bound (ICC = 0.98) suggests that case characteristics had minimal independent influence on response.

### Lawton-Young Supplementary Grade Exploration (Hemorrhage Status and Age)

Respondents indicated whether two LY components, hemorrhagic presentation and patient age (less than 20, 20–40, greater than 40 years; **Figure 2**), would influence their willingness to consider molecular therapy. Across all SM grades, providers consistently reported that prior rupture (OR, 2.44; 95% CI, 1.70–3.50; *P* < .001) and younger age (OR for age <20 years, 3.47; 95% CI, 2.26 –5.32; *P* < .001) increased their likelihood of considering both drug therapy and gene editing, with this pattern most pronounced at high SM grades (**eTable 3**). In subgroup analyses, the rupture effect was consistent for drug therapy (OR, 2.49; 95% CI, 1.73–3.58; *P* < .001) and gene editing (OR, 3.47; 95% CI, 2.14–5.63; *P* < .001). Pediatric age (<20 years) showed the strongest effect, followed by age 20– 40 (OR, 2.67; 95% CI, 1.88–3.79; *P* < .001) and age >40 (OR, 1.71; 95% CI, 1.19–2.47; *P* = 0.004) (**eTable 4**).

### Sensitivity Analyses

Primary findings were robust across all prespecified sensitivity analyses (**eTable 5**). Adjustment for provider training background yielded a similar effect size (S1: OR, 9.08; 95% CI, 3.75– 22.00), as did adjustment for AVM location (S2: OR, 16.01; 95% CI, 4.56–56.22). Restriction to complete cases produced comparable results (S3: OR, 10.37; 95% CI, 4.09–26.30), and dichotomization of SM grade confirmed the direction and significance of the association (S4: ORs 6.63–25.78; all *P* < .001).

### Implementation Concerns

Respondents identified several potential barriers to clinical adoption of molecular targeted therapies (**eTable 6**). The most frequently cited concerns for drug therapy were potential side effects: 33/48 (69%), patient safety: 29/48 (60%), and insufficient preclinical data: 28/48 (58%). Cost was a more prominent concern for gene editing: 21/48 (44%) than for drug therapy: 15/48 (31%).

## Discussion

Current knowledge in the literature indicates the necessity for exploration of the potential utility of newly discovered drug and gene editing therapy in bAVMs. For example, the *KRAS^G12C^*inhibitor sotorasib produced significant regression of extracranial *KRAS^G12C^*-driven facial vascular malformations in 2 adult patients and ameliorated bAVMs in mice^7^. The MEK inhibitor trametinib normalized cerebrovascular architecture in *KRAS*-mutant bAVM mouse models^8–10^ and stabilized disease in 2 of 3 human bAVM patients with unknown genotype, although 1 patient ruptured during therapy^11^. An ongoing phase II trial is assessing adjuvant trametinib in genotype-agnostic brain AVMs (NCT06098872). Finally, CRISPR-Cas mediated suppression of *Kras* repressed bAVMs in a *KRAS^G12D^* mouse model^12^. Therefore, moving forward by assessing expert clinical opinions regarding which factors push toward when and how to integrate these potentially breakthrough precision medical therapies into practice via fine-tuning the approach to clinical trials is an appropriate – and necessary – next step.

In this survey of 48 multidisciplinary cerebrovascular providers, broad willingness to consider precision medicine therapies for bAVMs was identified, with drug or gene editing therapy endorsement increasing more than 10-fold per unit increase in SM grade (OR, 10.41; 95% CI, 4.10– 26.42). This dose-response relationship aligns with clinical intuition: as bAVM complexity increases, conventional treatment options narrow and providers become increasingly receptive to novel paradigms. Notably, providers shifted from salvage to first-line endorsement with increasing SM grade, suggesting that specialists view molecular therapy not merely as a last resort but as a potentially disease-modifying strategy for complex lesions. The independent effects of rupture status and pediatric age further indicate that providers are already reasoning beyond anatomy alone, integrating clinical presentation and patient-level variables. These attitudes align with broader regulatory trends toward expediting trials for rare, genetically defined disorders, including FDA breakthrough therapy and rare pediatric disease frameworks^16^.

One question is whether existing clinical and preclinical evidence justifies precision medicine approaches in bAVM patients. While near-unanimous drug therapy feasibility endorsement (90%) and trial support (92%) are directionally consistent with the evidence trajectory, they outpace the current human evidence base. For example, sotorasib produced significant regression of extracranial *KRAS*^G12C^-driven AVMs in two adults and bAVMs in mice^7^. Three pediatric bAVM patients were treated with trametinib, of whom two demonstrated stable disease while one hemorrhaged during therapy^11^. Importantly, however, bAVM mutation status was unknown. The ongoing phase II trial of genotype-agnostic adjuvant trametinib in brain AVMs undergoing resection (NCT06098872) represents a critical first step towards implementing medical therapies. Although our results revealed independent effects of rupture status and pediatric age, preclinical models have not incorporated rupture state or developmental context, highlighting a translational gap. One proof-of-concept study demonstrated that CRISPR-Cas9 mediated prevention of *Kras* ameliorated KRAS^G12D^-driven bAVMs in mice^12^, though long-term safety, a translational delivery method and independent replication require further study.

A genotype-anchored framework is likely to provide the most robust evidence for precision bAVM treatment^13^. Implementing this approach requires an endovascular-enabled molecular diagnostic framework for variant detection and therapeutic monitoring by endoluminal and/or liquid biopsy^17–19^. Our data indicate universal willingness (100%) to implement endovascular-enabled molecular diagnostic strategies into clinical workstreams, but only a fraction of stakeholders have performed endoluminal (28%) or liquid (20%) biopsy for bAVMs. This discrepancy may reflect limited availability of validated diagnostic platforms, although unfamiliarity with technique and institutional infrastructure cannot be excluded. Prospective validation, including sensitivity thresholds for detecting variants at ultra-low allele frequency (<4%), is essential to enable the genotype-directed patient selection that respondents’ attitudes presuppose.

The SM grading scale comprises 12 distinct anatomical-angioarchitectural combinations that collapse into five ordinal grades, with substantial heterogeneity within intermediate grades – particularly grades III and IV^14^. To capture bAVM angioarchitectural and phenotypic variability, we proposed that an expanded taxonomy (**Figure 5**) is required to subtype bAVMs, determine candidacy for clinical trials, and evaluate treatment response. This study’s findings support construction of a revised bAVM genomic-phenomic taxonomy—incorporating genotype, angioarchitectural features, and clinical features—to redefine bAVMs as a molecularly tractable disease^13^. These data suggest clinical trials should prioritize higher-grade bAVMs flanking the boundaries of operability^20^. Incorporating genotype stratification, systematized neuroanatomical classification, and consensus endpoints beyond volumetric reduction (e.g., biomarker response, angioarchitectural remodeling) clinical trials may yield actionable findings. The lower enthusiasm displayed for gene editing (52% vs. 83% for drug therapy) appropriately reflects its earlier translational stage and higher costs, but these attitudes will likely shift as techniques mature. Multidisciplinary adjudication, endorsed by >80% of respondents, should be a cornerstone of any clinical trial framework.

**Figure 5.**
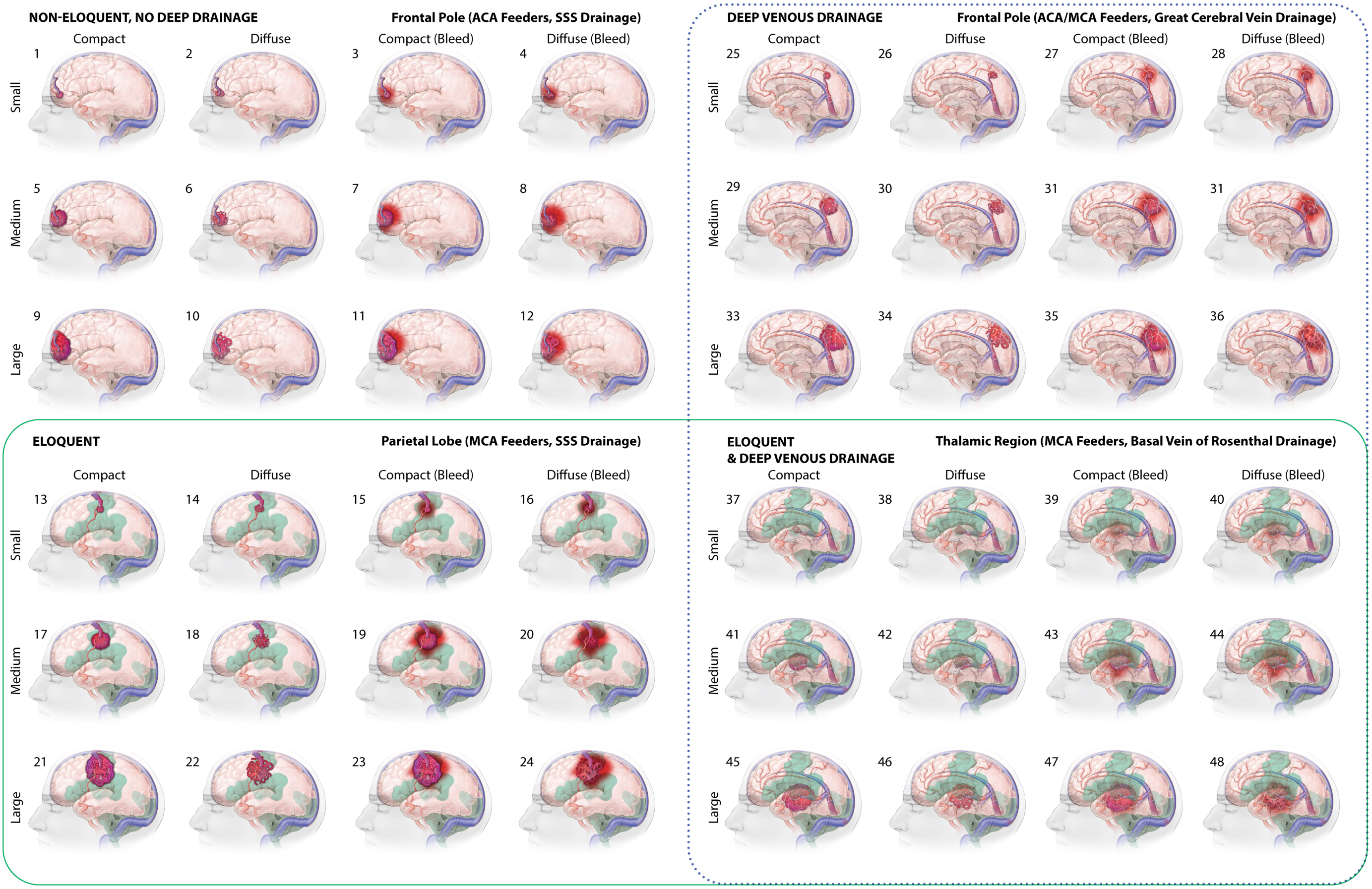
Expanded AVM taxonomy to guide clinical trial design for testing precision medicine therapeutics. This schematic stratifies bAVMs by anatomical location, venous drainage pattern, nidus size, and eloquence, illustrating the phenotypic heterogeneity within individual SM grades, particularly grades III and IV, that is not captured by ordinal grading alone. Green shading (with green box surrounding) highlights bAVM subtypes in eloquent locations; dotted-blue line indicates bAVMs that have deep venous drainage, a feature associated with increased bAVM rupture risk. Neurosurgical or radiosurgical intervention for bAVMs in eloquent location with deep venous draining (lower right block) represent the target population for precision medicine trial enrollment. Illustration by Laura Roy Illustrations.

Our study has several potential limitations. Hypothetical case vignettes do not fully capture the heterogeneity of bAVMs in clinical practice or nuances of patient preference. The instrument was developed de novo and has not been externally validated, and social desirability bias may have inflated endorsement rates. The high ICCs (0.70-0.98) indicate that most variance was attributable to stable inter-provider differences rather than case-level features, suggesting that individual provider philosophy may disproportionately influence therapy endorsement, a factor warranting consideration in future trial design. Survey distribution through professional networks to elicit responders introduces potential selection bias toward academic providers, although the participants are authors of approximately 2,175 AVM-related peer-reviewed publications, thereby proven to be clinical science experts whose opinions are worth eliciting. Finally, molecularly targeted therapy for bAVMs remains investigational, and these findings capture attitude at a single time point that will evolve as evidence accumulates.

Overall, cerebrovascular providers demonstrate broad but measured willingness to consider these targeted therapies for bAVMs, with strongest endorsement for drug treatment of high-grade lesions where conventional options are limited. The convergence of cerebrovascular genetics, preclinical therapeutic models, emerging endovascular enabled molecular diagnostic strategies, and now documented provider readiness creates a compelling rationale for genotype-stratified clinical trials evaluating precision medicine treatments for bAVMs.

## Previous presentation

None

## Previous publication

None

## Data Availability

All data produced in the present study are available upon reasonable request to the authors.

## SUPPLEMENTARY TABLES

**eTable 1.**
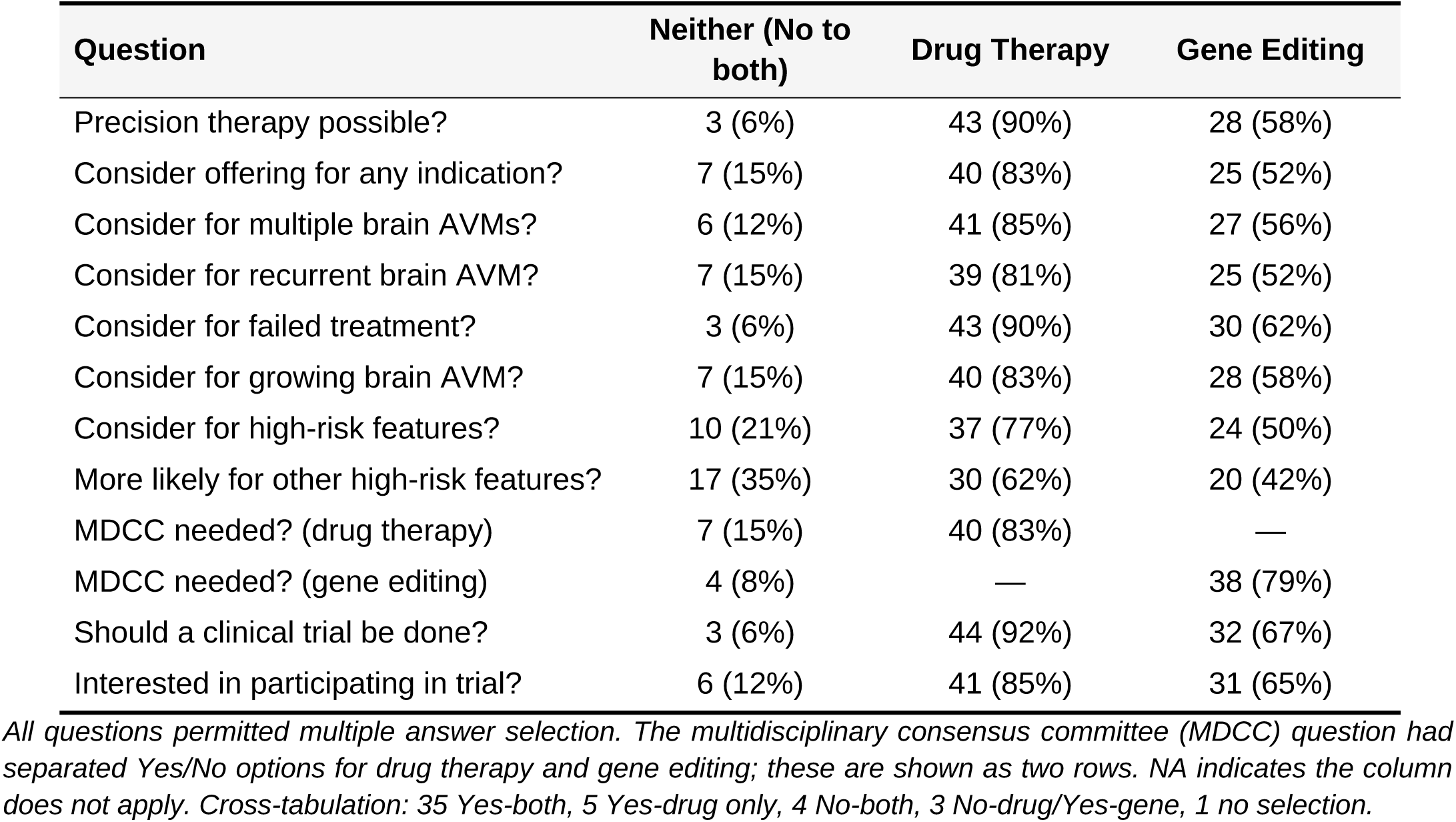
Provider attitudes towards drug and gene editing therapies for bAVMs (N = 48). Respondents could select multiple options per question. Values are n (%). The multidisciplinary committee question used separate Yes/No response options for drug therapy and gene editing, and is shown as two separate rows. Abbreviations include multidisciplinary consensus committee (MDCC).

| **Question** | **Neither (No to both)** | **Drug Therapy** | **Gene Editing** |
|---|---|---|---|
| Precision therapy possible? | 3 (6%) | 43 (90%) | 28 (58%) |
| Consider offering for any indication? | 7 (15%) | 40 (83%) | 25 (52%) |
| Consider for multiple brain AVMs? | 6 (12%) | 41 (85%) | 27 (56%) |
| Consider for recurrent brain AVM? | 7 (15%) | 39 (81%) | 25 (52%) |
| Consider for failed treatment? | 3 (6%) | 43 (90%) | 30 (62%) |
| Consider for growing brain AVM? | 7 (15%) | 40 (83%) | 28 (58%) |
| Consider for high-risk features? | 10 (21%) | 37 (77%) | 24 (50%) |
| More likely for other high-risk features? | 17 (35%) | 30 (62%) | 20 (42%) |
| MDCC needed? (drug therapy) | 7 (15%) | 40 (83%) | — |
| MDCC needed? (gene editing) | 4 (8%) | — | 38 (79%) |
| Should a clinical trial be done? | 3 (6%) | 44 (92%) | 32 (67%) |
| Interested in participating in trial? | 6 (12%) | 41 (85%) | 31 (65%) |

*All questions permitted multiple answer selection. The multidisciplinary consensus committee (MDCC) question had separated Yes/No options for drug therapy and gene editing; these are shown as two rows. NA indicates the column does not apply. Cross-tabulation: 35 Yes-both, 5 Yes-drug only, 4 No-both, 3 No-drug/Yes-gene, 1 no selection.*

**eTable 2.**
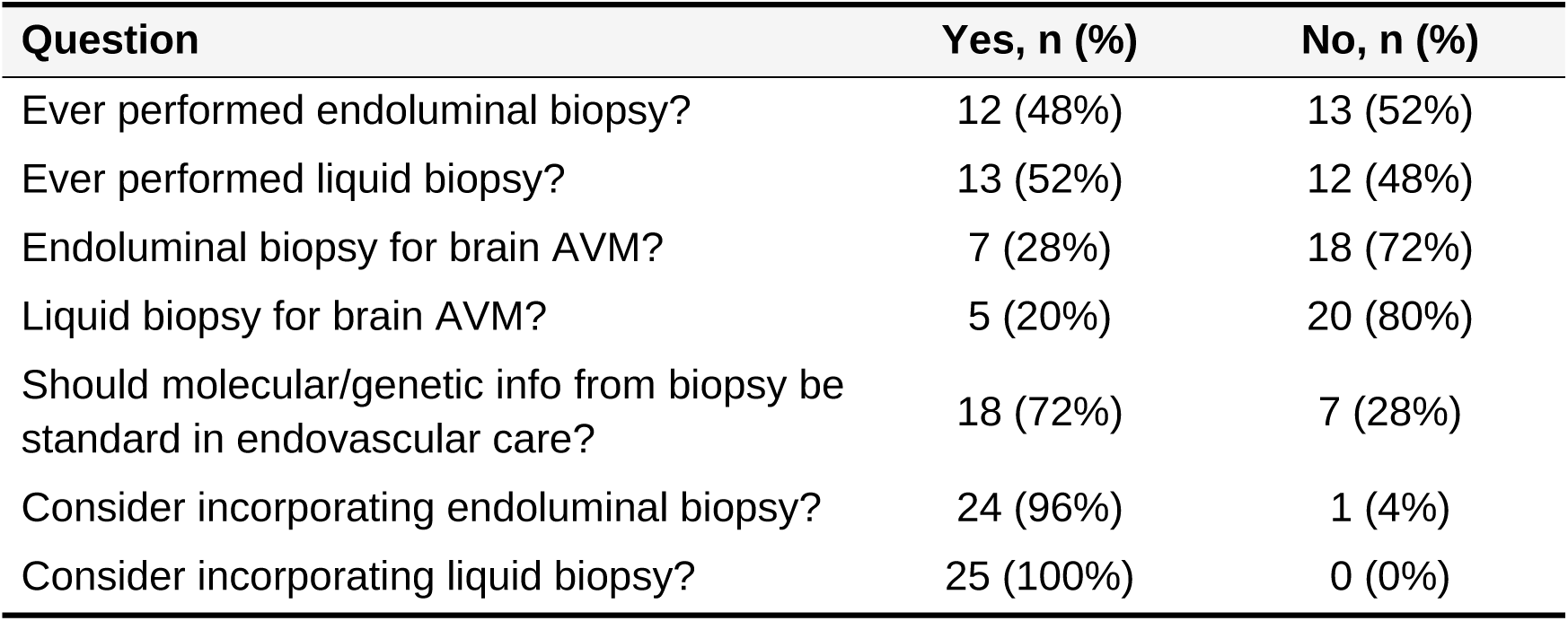
Endovascular provider attitudes toward molecular diagnostic techniques for bAVMs (n = 25). 25 of 26 respondents with endovascular training. 1 was excluded due to missing training response question. Values are n (%). Questions are check-all-that-apply where applicable.

| **Question** | **Yes, n (%)** | **No, n (%)** |
|---|---|---|
| Ever performed endoluminal biopsy? | 12 (48%) | 13 (52%) |
| Ever performed liquid biopsy? | 13 (52%) | 12 (48%) |
| Endoluminal biopsy for brain AVM? | 7 (28%) | 18 (72%) |
| Liquid biopsy for brain AVM? | 5 (20%) | 20 (80%) |
| Should molecular/genetic info from biopsy be standard in endovascular care? | 18 (72%) | 7 (28%) |
| Consider incorporating endoluminal biopsy? | 24 (96%) | 1 (4%) |
| Consider incorporating liquid biopsy? | 25 (100%) | 0 (0%) |

**eTable 3.** Provider endorsement of molecular targeted therapy across 12 bAVM case vignettes (Figure 2) by Spetzler-Martin grade, stratified by treatment timing (first line), hemorrhagic presentation, and patient age (< 20, 20-40, and > 40 years). All questions used a check-all-that-apply format. Values are counts (denominators in column header row). Note that nidus compactness (third Lawton-Young component) was not assessed in this survey.

| **bAVM case vignette (Figure 2)** | **A** | **B** | **C** | **D** | **E** | **F** | **G** | **H** | **I** | **J** | **K** | **L** |
|---|---|---|---|---|---|---|---|---|---|---|---|---|
| **SM grade** | **I** | **III** | **II** | **IV** | **II** | **IV** | **III** | **V** | **I** | **II** | **III** | **IV** |
| **Q1: If this bAVM had an ‘actionable’ mutation, what molecular targeted therapy options would you consider?** |  |  |  |  |  |  |  |  |  |  |  |  |
| None | 12 | 5 | 6 | 3 | 9 | 4 | 5 | 3 | 11 | 6 | 7 | 3 |
| Drug therapy – 1st line | 5 | 11 | 17 | 26 | 7 | 21 | 21 | 27 | 6 | 12 | 14 | 21 |
| Gene editing – 1st line | 4 | 8 | 10 | 15 | 6 | 12 | 14 | 16 | 4 | 6 | 8 | 13 |
| Drug therapy – Adjuvant | 7 | 13 | 8 | 11 | 8 | 15 | 12 | 13 | 8 | 11 | 15 | 15 |
| Gene editing – Adjuvant | 6 | 10 | 6 | 8 | 5 | 12 | 6 | 10 | 5 | 9 | 12 | 13 |
| Drug therapy – Salvage | 23 | 19 | 15 | 8 | 22 | 14 | 14 | 10 | 18 | 19 | 16 | 13 |
| Gene editing – Salvage | 16 | 14 | 9 | 5 | 15 | 11 | 10 | 8 | 13 | 15 | 12 | 10 |
| **Q2: Would a prior rupture influence your decision to offer molecular targeted therapy?** |  |  |  |  |  |  |  |  |  |  |  |  |
| No influence | 21 | 20 | 18 | 20 | 22 | 22 | 19 | 20 | 19 | 21 | 18 | 17 |
| More likely – Drug | 5 | 7 | 9 | 12 | 5 | 7 | 13 | 15 | 7 | 7 | 9 | 15 |
| Less likely – Drug | 12 | 10 | 9 | 5 | 9 | 8 | 5 | 1 | 10 | 9 | 9 | 4 |
| More likely – Gene | 2 | 4 | 7 | 7 | 2 | 5 | 9 | 12 | 3 | 5 | 7 | 11 |
| Less likely – Gene | 8 | 8 | 8 | 1 | 6 | 5 | 2 | 1 | 7 | 6 | 5 | 3 |
| **Q3a: If this was a pediatric patient (<20 years old), would that influence your decision to administer molecular targeted therapy?** |  |  |  |  |  |  |  |  |  |  |  |  |
| No influence | 24 | 25 | 24 | 20 | 23 | 23 | 20 | 20 | 23 | 24 | 21 | 20 |
| More likely – Drug | 5 | 6 | 7 | 14 | 4 | 10 | 12 | 14 | 4 | 7 | 8 | 13 |
| Less likely – Drug | 8 | 5 | 4 | 3 | 8 | 3 | 3 | 2 | 8 | 4 | 5 | 1 |
| More likely – Gene | 3 | 4 | 5 | 10 | 2 | 7 | 9 | 10 | 3 | 5 | 9 | 10 |
| Less likely – Gene | 7 | 4 | 3 | 2 | 7 | 3 | 2 | 1 | 5 | 2 | 2 | 1 |
| **Q3b: If this was a patient between 20–40 years old, would that influence your decision to administer molecular targeted therapy?** |  |  |  |  |  |  |  |  |  |  |  |  |
| No influence | 28 | 30 | 27 | 22 | 27 | 26 | 22 | 21 | 28 | 28 | 26 | 20 |
| More likely – Drug | 5 | 2 | 5 | 13 | 3 | 7 | 10 | 13 | 2 | 4 | 4 | 12 |
| Less likely – Drug | 5 | 4 | 3 | 2 | 5 | 3 | 3 | 2 | 5 | 4 | 4 | 2 |
| More likely – Gene | 3 | 2 | 5 | 9 | 2 | 5 | 7 | 9 | 1 | 1 | 4 | 9 |
| Less likely – Gene | 4 | 3 | 2 | 2 | 4 | 2 | 1 | 1 | 3 | 1 | 3 | 2 |
| **Q3c: If this was a patient >40 years old, would that influence your decision to administer molecular targeted therapy?** |  |  |  |  |  |  |  |  |  |  |  |  |
| No influence | 28 | 28 | 25 | 24 | 28 | 28 | 26 | 24 | 28 | 28 | 28 | 24 |
| More likely – Drug | 9 | 6 | 10 | 11 | 7 | 6 | 9 | 11 | 5 | 6 | 6 | 10 |
| Less likely – Drug | 2 | 3 | 1 | 1 | 1 | 1 | 1 | 1 | 3 | 2 | 3 | 2 |
| More likely – Gene | 4 | 2 | 6 | 5 | 2 | 2 | 3 | 6 | 1 | 1 | 0 | 4 |
| Less likely – Gene | 1 | 2 | 0 | 0 | 0 | 1 | 0 | 0 | 1 | 0 | 1 | 0 |

**eTable 4.** Mixed-effects logistic regression results for 10 binary therapy endorsement outcomes. Each model estimated the odds ratio (OR) with 95% confidence interval (CI) per Spetzler-Martin grade unit increase, with a random intercept for respondent to account for repeated-measures. All *P* values remained significant after Benjamini-Hochberg false discovery rate (FDR) (adjusted P < .001 for all outcomes).

| Group | Outcome | Events | OR (95% CI) | *P* Value | ICC |
|---|---|---|---|---|---|
| Primary | Any molecular therapy | 364 | 10.41 (4.10–26.42) | <0.001 | 0.98 |
| Primary | Any drug therapy | 363 | 6.74 (3.21–14.14) | <0.001 | 0.98 |
| Primary | Any gene editing | 244 | 2.51 (1.61–3.92) | <0.001 | 0.94 |
| Primary | 1st-line drug | 188 | 4.21 (2.98–5.96) | <0.001 | 0.77 |
| Primary | 1st-line gene editing | 116 | 3.38 (2.28–5.00) | <0.001 | 0.88 |
| Secondary | None (no therapy) | 74 | 0.14 (0.07–0.30) | <0.001 | 0.97 |
| Secondary | Adjuvant drug | 136 | 1.66 (1.29–2.14) | <0.001 | 0.76 |
| Secondary | Adjuvant gene editing | 102 | 1.59 (1.23–2.06) | <0.001 | 0.72 |
| Secondary | Salvage drug | 191 | 0.53 (0.42–0.67) | <0.001 | 0.74 |
| Secondary | Salvage gene editing | 138 | 0.67 (0.52–0.85) | <0.001 | 0.70 |

**eTable 5.**
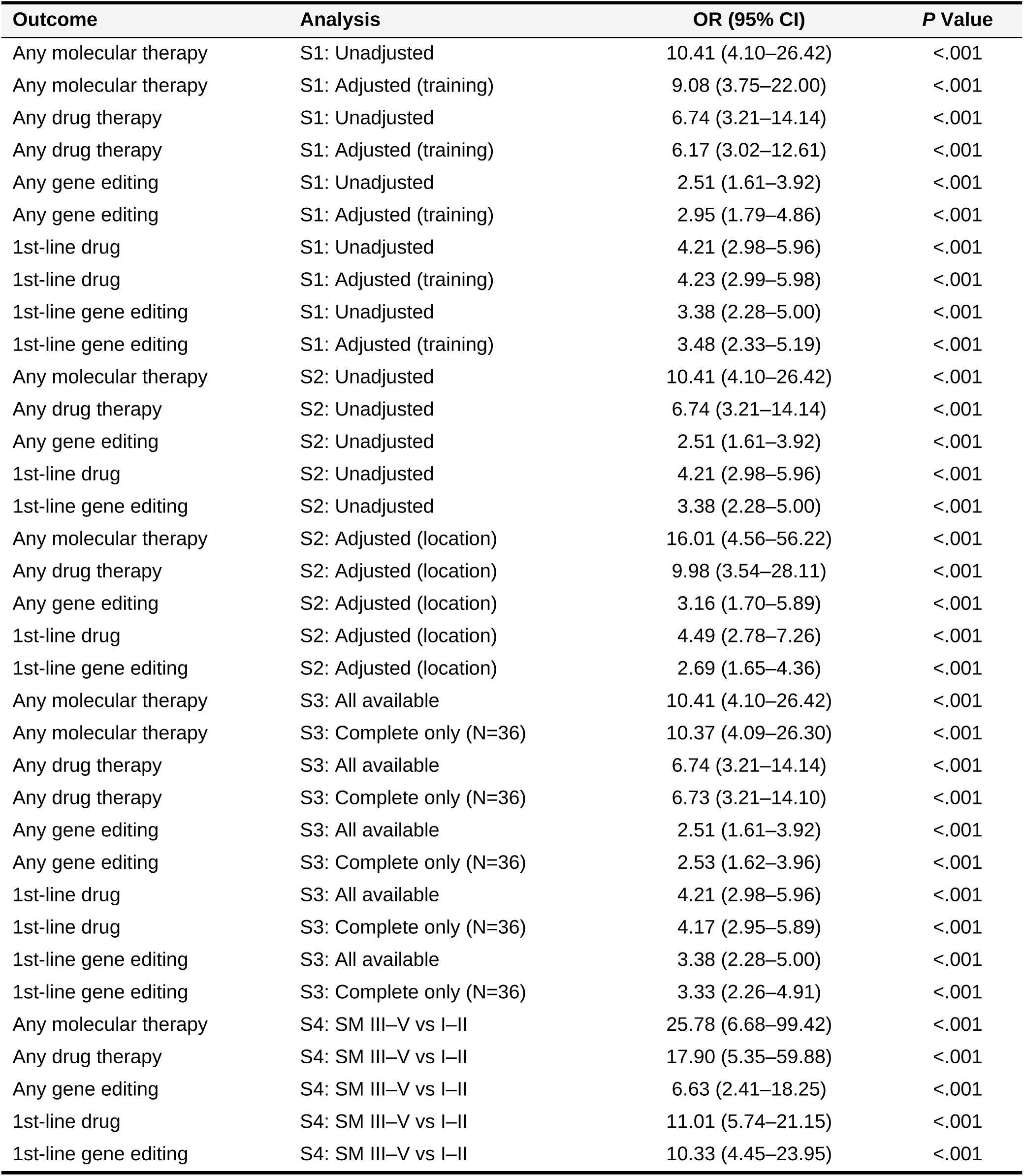
Sensitivity analyses for the five primary mixed-effects logistic regression outcomes. S1: adjustment for provider training subspecialty (fixed effect). S2: adjustment for bAVM anatomical location (fixed effect). S3: model restricted to respondents who completed all 12 scenarios. S4: dichotomization of SM grade (III–V vs I–II). All *P* values < .001 after Benjamini-Hochberg FDR correction.

**eTable 6.**
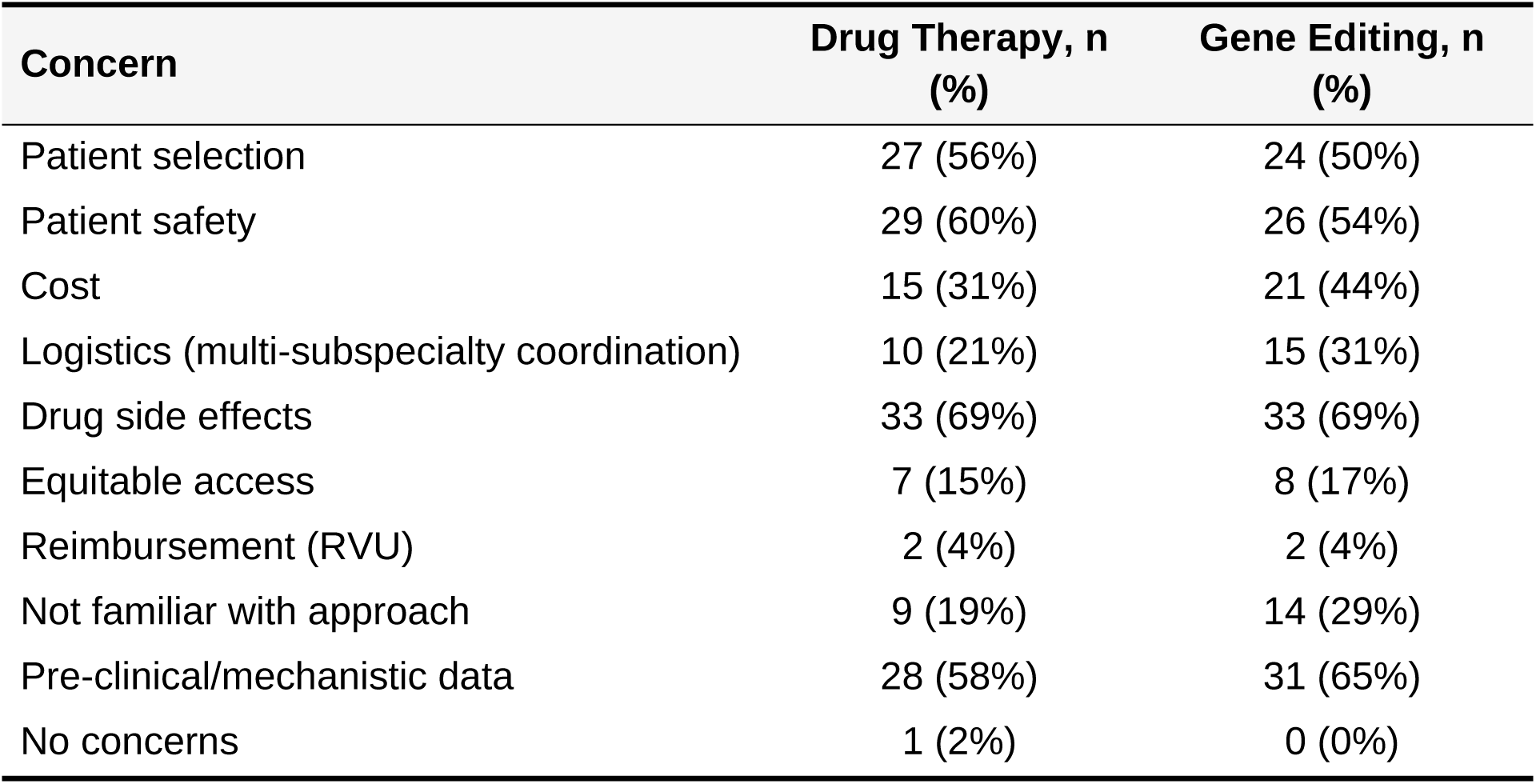
Provider barriers to clinical implementation for drug and gene editing therapies for bAVMs (n = 48 respondents). Respondents could select multiple concerns. Values are N (%).

| **Concern** | **Drug Therapy, n (%)** | **Gene Editing, n (%)** |
|---|---|---|
| Patient selection | 27 (56%) | 24 (50%) |
| Patient safety | 29 (60%) | 26 (54%) |
| Cost | 15 (31%) | 21 (44%) |
| Logistics (multi-subspecialty coordination) | 10 (21%) | 15 (31%) |
| Drug side effects | 33 (69%) | 33 (69%) |
| Equitable access | 7 (15%) | 8 (17%) |
| Reimbursement (RVU) | 2 (4%) | 2 (4%) |
| Not familiar with approach | 9 (19%) | 14 (29%) |
| Pre-clinical/mechanistic data | 28 (58%) | 31 (65%) |
| No concerns | 1 (2%) | 0 (0%) |

## SUPPLEMENTARY FIGURES

**eFigure 1.**
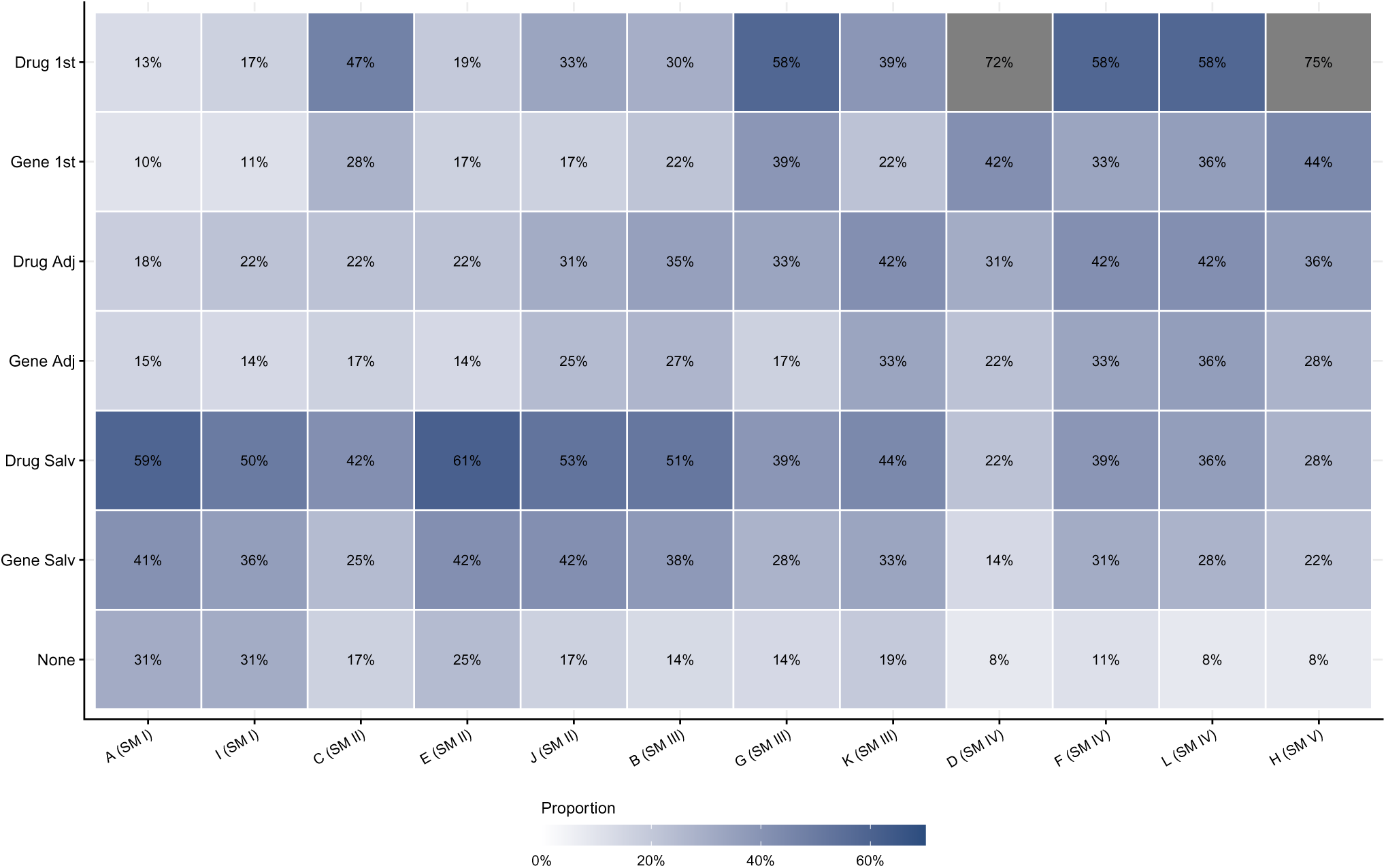
Treatment endorsement heatmap across 12 bAVM case vignettes. X-axis: cases sorted by ascending SM grade. Y-axis: treatment-reliance ordinal scale reflecting increasing clinical commitment to molecular therapy (None → Salvage → Adjuvant → 1st-line). Cell values show the proportion of respondents endorsing each treatment option. Case designation (x-axis) corresponds to panel in Figure 2.

**eFigure 2.**
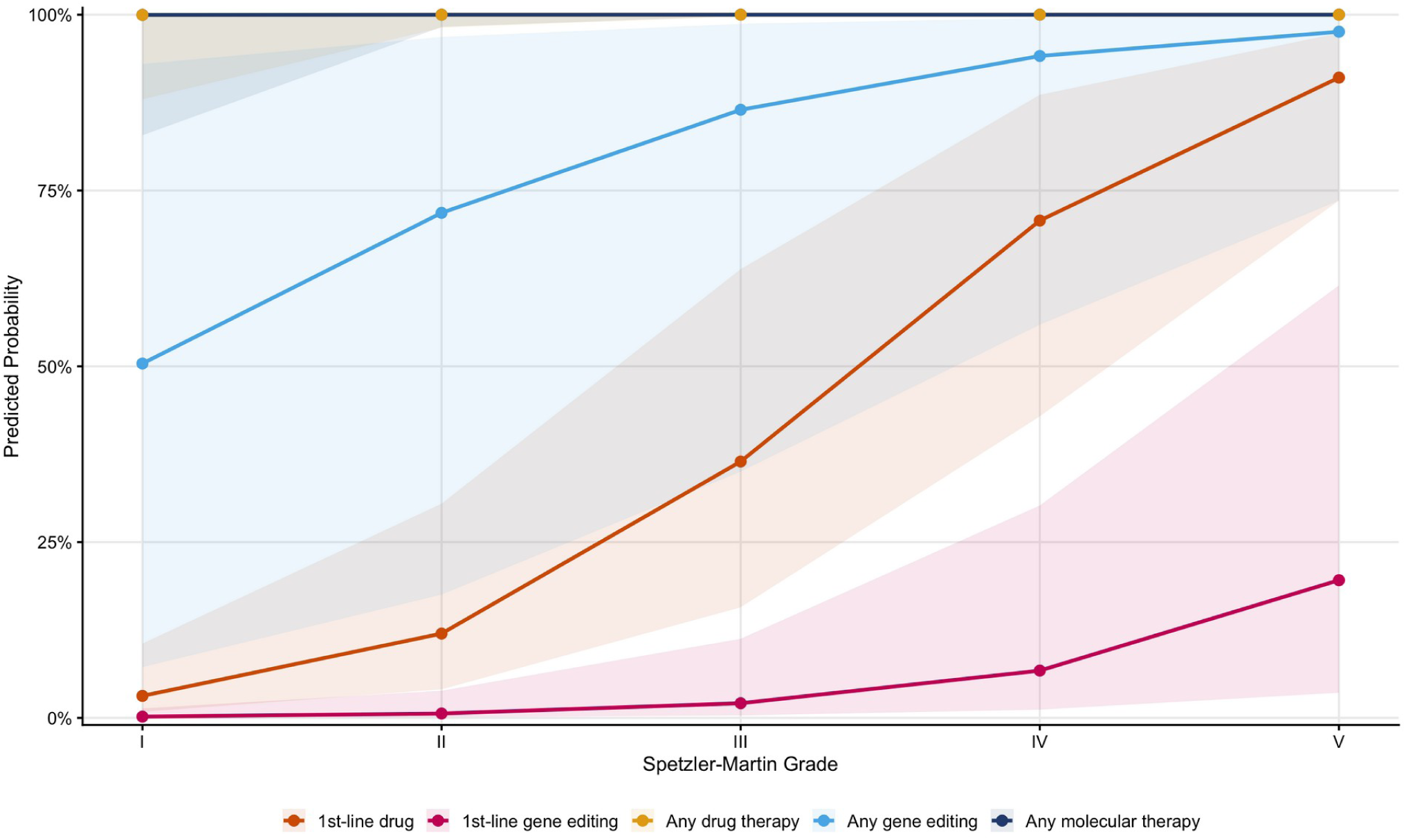
Observed proportions of therapy endorsement by Spetzler–Martin grade with exact binomial 95% confidence intervals. Same data as Figure 4 with CI ribbons.

## SUPPLEMENTARY NOTE 1

**Supplementary Note 1.** Survey Instrument. The following questions were administered via REDCap. Response options are listed below each question. Questions permitting multiple selections are noted. Case scenario questions (Cases 1–12) shared an identical question template and are shown once with the per-case anatomical description listed separately.

## Section A: Demographics

1. Have you read ‘An opportune time for targeted brain arteriovenous malformation therapy’ by Hale et al. (2025) published in Journal of Neurosurgery? *Response options: No; Yes*
2. What is your age? Response options: 25–34; 35–44; 45–54; 55–64; 65–74; > 75
3. What is your sex? *Response options: Male; Female; Prefer not to answer*
4. Are you US-based or non-US based? *Response options: US-Based; Non-US based*
5. What is your practice/training background? *Response options: Adult – Open & Endovascular; Adult – Open vascular only; Adult – Endovascular only (via neurology, radiology); Pediatric – Performs open vascular cases (no dedicated open vascular fellowship); Pediatric – Completed both pediatric and open vascular fellowships; Pediatric – Endovascular alone (via neurology, radiology)*
6. What is your practice type? *Response options: Adult only; Pediatrics only; Combined adult & pediatric (majority adult); Combined adult & pediatric (majority pediatrics)*
7. What is your practice setting? *Response options: Adults-only hospital; Stand-alone children’s hospital; Combined adult-pediatric hospital; Other*
8. Are you board-certified? *Response options: No; No, I am board eligible; Yes*
9. What is your academic rank? *Response options: Instructor; Assistant Professor; Associate Professor; Full Professor; Not applicable*
10. What is your highest level of research training? *Response options: No formal research training; Funded research fellowship non-degree (minimum 1 year); Master’s (MS, MSc, MPH, etc.); Doctorate (PhD); Other*
11. Have you ever been awarded a research grant (check all that apply)? *Response options: No; Yes – NIH (or equivalent) as PI; Yes – NIH (or equivalent) as co-I; Yes – Foundation grant as PI; Yes – Foundation grant as co-I; Yes – Institutional award as PI; Yes – Institutional award as co-I*
12. Have you heard the term ‘endoluminal, coil, or endothelial’ biopsy before reading the key definitions? *Response options: No; Yes*
13. Have you heard the term ‘liquid’ biopsy before reading the key definitions? *Response options: No; Yes*

## Section B: Drug Therapy for bAVMs

*All questions in this section are check-all-that-apply*.

1. Do you think that molecular targeted therapy for select bAVMs is possible? *Response options: No; Yes – drug therapy; Yes – gene editing*
2. Would you consider offering molecular targeted therapy for brain AVMs for any indication? *Response options: No; Yes – drug therapy; Yes – gene editing*
3. Would you consider offering molecular targeted therapy for a patient with multiple bAVMs? *Response options: No; Yes – drug therapy; Yes – gene editing*
4. Would you consider offering molecular targeted therapy for a bAVM that recurred after treatment? *Response options: No; Yes – drug therapy; Yes – gene editing*
5. Would you consider offering molecular targeted therapy for a bAVM that failed other treatment modalities? *Response options: No; Yes – drug therapy; Yes – gene editing*
6. Would you consider offering molecular targeted therapy for a bAVM that was growing over time? *Response options: No; Yes – drug therapy; Yes – gene editing*
7. Would you consider offering molecular targeted therapy for a bAVM that harbored a high-risk feature that was treated (i.e., intranidal aneurysm)? *Response options: No; Yes – drug therapy; Yes – gene editing*
8. Would you be more likely to offer molecular targeted therapy for a bAVM that harbored other high-risk features (venous varix, venous outflow stenosis, flow-related aneurysm, etc.)? *Response options: No; Yes – drug therapy; Yes – gene editing*
9. Is a multidisciplinary adjudication/consensus committee needed to determine which bAVM patients should receive molecular targeted therapy? *Response options: No – drug therapy; Yes – drug therapy; No – gene editing; Yes – gene editing*
10. Should a randomized clinical trial of molecular targeted therapy for bAVMs be performed? *Response options: No; Yes – drug therapy; Yes – gene editing*
11. Are you interested in participating in a randomized clinical trial for molecular targeted therapy for bAVMs? *Response options: No; Yes – drug therapy; Yes – gene editing*
12. Have you treated a bAVM patient with drug therapy? *Response options: No; Yes*
13. Have you treated an extracranial AVM patient with drug therapy? *Response options: No; Yes*

## Section C: Endovascular-Specific Questions

*Displayed only to respondents with endovascular training*.

1. Are you endovascular trained? *Response options: Yes; No*
2. Have you ever performed ‘endoluminal’ biopsy for any indication? *Response options: No; Yes*
3. Have you ever performed ‘liquid’ biopsy for any indication? *Response options: No; Yes*
4. Have you ever performed ‘endoluminal’ biopsy for a patient with a brain AVM? *Response options: No; Yes*
5. Have you ever performed ‘liquid’ biopsy for a patient with a brain AVM? *Response options: No; Yes*
6. Should molecular/genetic information obtained from ‘endoluminal’ and/or ‘liquid’ biopsy become a standard component of endovascular care? *Response options: No; Yes*
7. Would you consider incorporating ‘endoluminal’ biopsy into your practice? *Response options: No; Yes*
8. Would you consider incorporating ‘liquid’ biopsy into your practice? *Response options: No; Yes*

## Section D: Case Scenarios (Cases 1–12)

Each case presents a specific brain AVM with defined Spetzler–Martin grade, location, size, arterial feeders, and venous drainage. For each case, the following three questions were asked (all check-all-that-apply):

**Q1 (Initial Treatment):** If this bAVM had an ‘actionable’ mutation, what molecular targeted therapy options would you consider?

*Response options: None; Drug therapy – 1st line; Gene editing – 1st line; Drug therapy – Adjuvant with embo/XRT; Gene editing – Adjuvant with embo/XRT; Drug therapy – Salvage after failed surgery/XRT/embo; Gene editing – Salvage after failed surgery/XRT/embo*

**Q2 (Rupture History):** Would a prior rupture influence your decision to offer molecular targeted therapy?

*Response options: No; Yes – more likely to consider drug therapy; Yes – less likely to consider drug therapy; Yes – more likely to consider gene editing therapy; Yes – less likely to consider gene editing therapy*

**Q3 (Age Consideration):** If this was a [pediatric patient < 20 years old / patient between 20–40 years old / patient > 40 years old], would that influence your decision to administer molecular targeted therapy?

*Response options: No; Yes – more likely to consider drug therapy; Yes – less likely to consider drug therapy; Yes – more likely to consider gene editing therapy; Yes – less likely to consider gene editing therapy*

*(Asked separately for each of the three age categories.)*

### Case Descriptions

Case 1: SM grade I AVM in the frontal pole, < 3 cm, with ACA feeders, and superficial drainage to the superior sagittal sinus.

Case 2: SM grade III AVM in the frontal pole, > 6 cm in size, with ACA feeders, and superficial drainage to the superior sagittal sinus.

Case 3: SM grade II AVM in the eloquent motor/somatosensory cortex, < 3 cm in size, with MCA feeders, and superficial drainage to the superior sagittal sinus.

Case 4: SM grade IV AVM in the eloquent motor/somatosensory cortex, > 6 cm in size, with MCA feeders, and superficial drainage to the superior sagittal sinus.

Case 5: SM grade II AVM in the non-eloquent precuneus, < 3 cm in size, with ACA & MCA feeders, and deep venous drainage.

Case 6: SM grade IV AVM in the non-eloquent precuneus, > 6 cm in size, with ACA & MCA feeders, and deep venous drainage.

Case 7: SM grade III AVM in the eloquent thalamus/basal ganglia, < 3 cm in size, with MCA perforator feeders, and deep venous drainage.

Case 8: SM grade V AVM in the eloquent thalamus/basal ganglia, > 6 cm in size, with MCA perforator feeders, and deep venous drainage.

Case 9: SM grade I AVM in the non-eloquent cerebellar hemisphere, < 3 cm in size, with PICA perforators, and superficial venous drainage.

Case 10: SM grade II AVM in the non-eloquent cerebellar hemisphere, 3–6 cm in size, with PICA perforators, and superficial venous drainage.

Case 11: SM grade III AVM in the eloquent vermis, < 3 cm in size, with PICA perforators, and deep venous drainage.

Case 12: SM grade IV AVM in the eloquent vermis, 3–6 cm in size, with PICA perforators, and deep venous drainage.

## Section E: Concerns

*All questions are check-all-that-apply*.

1. What are your general concerns (if any) for implementing drug therapy for patients with bAVMs? Response options: Patient selection; Patient safety (risk of rupture or stroke whilst on therapy compared to natural history); Cost; Logistics → multi-subspecialty coordination; Drug side effects; Equitable access for patients; Reimbursement (RVU); Not familiar with approach; Pre-clinical/mechanistic data in support of drug therapy; No concerns
2. What are your general concerns (if any) for implementing gene editing therapy for patients with bAVMs? Response options: Patient selection; Patient safety (risk of rupture or stroke whilst on therapy compared to natural history); Cost; Logistics → multi-subspecialty coordination; Gene editing side effects; Equitable access for patients; Reimbursement (RVU); Not familiar with technique/approach; Pre-clinical/mechanistic data in support of gene editing therapy; No concerns
3. What general concerns (if any) do you have performing endoluminal biopsy for bAVMs? Response options: Patient/lesion/vessel selection; Patient safety (risk of rupture or stroke); Cost; Logistics (i.e., operative time, new process, etc.); Discerning ‘clinically actionable’ information; Equitable access for patients; Reimbursement (RVU); Not familiar with technique/approach; No concerns
4. What general concerns (if any) do you have performing liquid biopsy for bAVMs? Response options: Patient/lesion/vessel selection; Patient safety (risk of rupture or stroke); Cost; Logistics (i.e., operative time, new process, etc.); Discerning ‘clinically actionable’ information; Equitable access for patients; Reimbursement (RVU); Not familiar with technique/approach; No concerns

## References

1. Nikolaev SI, Vetiska S, Bonilla X, et al. Somatic activating KRAS mutations in arteriovenous malformations of the brain. N Engl J Med. 2018;378(3):250–261. doi:10.1056/NEJMoa1709449

2. Hong T, Yan Y, Li J, et al. High prevalence of KRAS/BRAF somatic mutations in brain and spinal cord arteriovenous malformations. Brain. 2019;142(1):23–34. doi:10.1093/brain/awy307

3. Goss JA, Huang AY, Smith E, et al. Somatic mutations in intracranial arteriovenous malformations. PLoS One. 2019;14(12):e0226852. doi:10.1371/journal.pone.0226852

4. Priemer DS, Vortmeyer AO, Zhang S, Chang HY, Curless KL, Cheng L. Activating KRAS mutations in arteriovenous malformations of the brain: frequency and clinicopathologic correlation. Hum Pathol. 2019;89:33–39. doi:10.1016/j.humpath.2019.04.004

5. Gao S, Nelson J, Weinsheimer S, et al. Somatic mosaicism in the MAPK pathway in sporadic brain arteriovenous malformation and association with phenotype. J Neurosurg. 2022;136(1):148–155. doi:10.3171/2020.11.JNS202031

6. Hanrahan AJ, Chen Z, Rosen N, Solit DB. BRAF - a tumour-agnostic drug target with lineage-specific dependencies. Nat Rev Clin Oncol. 2024;21(3):224–247. doi:10.1038/s41571-023-00852-0

7. Fraissenon A, Bayard C, Morin G, et al. Sotorasib for vascular malformations associated with KRAS G12C mutation. N Engl J Med. 2024;391(4):334–342. doi:10.1056/NEJMoa2309160

8. Myint O, Freeman BH, Jeong J, et al. Trametinib decreased intracerebral hemorrhages and endothelial-to-mesenchymal transition in KRASG12V-induced brain arteriovenous malformations in mice. Stroke. 2026;57(3):779–793. doi:10.1161/STROKEAHA.125.052418

9. Park ES, Kim S, Huang S, et al. Selective endothelial hyperactivation of oncogenic KRAS induces brain arteriovenous malformations in mice. Ann Neurol. 2021;89(5):926–941. doi:10.1002/ana.26059

10. DeBose-Scarlett E, Ressler AK, Gallione CJ, et al. Somatic mutations in arteriovenous malformations in hereditary hemorrhagic telangiectasia support a bi-allelic two-hit mutation mechanism of pathogenesis. Am J Hum Genet. 2024;111(10):2283–2298. doi:10.1016/j.ajhg.2024.08.020

11. Suarez CF, Harb OA, Robledo A, et al. MEK signaling represents a viable therapeutic vulnerability of KRAS-driven somatic brain arteriovenous malformations. bioRxivorg. Published online May 15, 2024. doi:10.1101/2024.05.15.594335

12. Saito S, Nakamura Y, Miyashita S, et al. CRISPR/CasRx suppresses KRAS-induced brain arteriovenous malformation developed in postnatal brain endothelial cells in mice. JCI Insight. 2024;9(22). doi:10.1172/jci.insight.179729

13. Hale AT, Kalailingam P, Kundishora AJ, et al. An opportune time for targeted brain arteriovenous malformation therapy. J Neurosurg. 2025;143(6):1685–1688. doi:10.3171/2025.4.JNS25260

14. Spetzler RF, Martin NA. A proposed grading system for arteriovenous malformations. J Neurosurg. 1986;65(4):476–483. doi:10.3171/jns.1986.65.4.0476

15. Lawton MT, Kim H, McCulloch CE, Mikhak B, Young WL. A supplementary grading scale for selecting patients with brain arteriovenous malformations for surgery. Neurosurgery. 2010;66(4):702–713; discussion 713. doi:10.1227/01.NEU.0000367555.16733.E1

16. Prasad V, Makary MA. One pivotal trial, the new default option for FDA approval - ending the two-trial dogma. N Engl J Med. 2026;394(8):815–817. doi:10.1056/NEJMsb2517623

17. Winkler E, Wu D, Gil E, et al. Endoluminal biopsy for molecular profiling of human brain vascular malformations. Neurology. 2022;98(16):e1637–e1647. doi:10.1212/WNL.0000000000200109

18. Hale AT, Liu S, Huang F, et al. Endoluminal biopsy for vein of Galen malformation. Neurosurgery. 2024;95(5):1082–1088. doi:10.1227/neu.0000000000002986

19. Bhatia KD, Sylvester D, Padhye B, et al. Harnessing liquid biopsy to unveil RAS-MEK pathway somatic pathogenic variants in extracranial arterio-venous malformations. Commun Med (Lond). 2025;5(1):508. doi:10.1038/s43856-025-01174-1

20. Catapano JS, Frisoli FA, Nguyen CL, et al. Intermediate-grade brain arteriovenous malformations and the boundary of operability using the supplemented Spetzler-Martin grading system. J Neurosurg. 2022;136(1):125–133. doi:10.3171/2020.11.JNS203298

